# Monoclonal antibody dispensing during and around pregnancy: a descriptive analysis using electronic health records in Italy

**DOI:** 10.64898/2026.03.25.26349279

**Authors:** Elisabeth Aiton, Valeria Nazzari, Rosie Cornish, Benjamin G Faber, Christy Burden, Katherine Birchenall, Maria Carolina Borges, Deborah A Lawlor, Luisa Zuccolo

## Abstract

**Objective:** To describe trends in dispensing of monoclonal antibodies (mAbs) for autoimmune conditions during and around pregnancy.

**Design:** Descriptive study.

**Setting:** Lombardy, Italy between 2012 and 2024.

**Population:** All women of reproductive age (14-49 years) resident in Lombardy.

**Methods:** We described trends in mAb dispensations among women of reproductive age and the prevalence of mAb dispensing before, during and after pregnancy. We explored maternal factors associated with discontinuation.

**Main outcome measures:** Change in prescribing of mAbs over time in all women of reproductive age, and before, during and after pregnancy in those who became pregnant. Prevalence of discontinuation and switching mAbs around pregnancy.

**Results:** We included 3,049,175 women of reproductive age and 859,699 pregnancies. Prevalence of mAb dispensing during pregnancy increased over 60-fold over the study period, from 0.0041% (95%CI:0.00084, 0.012) in 2012 to 0.27% (95%CI:0.23, 0.32) in 2024. Pregnancy affected mAb dispensing, with mean prevalence decreasing from 0.080% (95%CI:0.074, 0.087) before pregnancy to 0.051% (95%CI:0.046, 0.057) by the third trimester. Over half (53.3%) of pre-existing users discontinued before or during pregnancy; discontinuation decreased over time, and varied substantially between mAbs. Switching mAbs during pregnancy was rare (3.3%). We found limited evidence that sociodemographic factors were associated with discontinuation, but that some health factors may be, such as use of assisted reproductive technology (OR=1.92, 95%CI:0.98-3.77).

**Conclusions:** Italian population-wide data from 2012-2024 show an increase in mAbs dispensed during pregnancy, and fewer instances of discontinuing these drugs over time. This may reflect recent changes in prescribing guidelines for mAbs in pregnancy.

## Introduction

In Europe, autoimmune conditions affect 7-13% of women, and are frequently diagnosed during reproductive age [1,2]. Moderate to severe autoimmune conditions are increasingly managed by monoclonal antibody therapeutics (mAbs) in high-income countries [3–5]. A recent Italian study found that 10.5% of women with psoriasis and 20.1% of women with rheumatoid arthritis used mAbs prior to pregnancy [6].

mAbs provide a targeted and effective treatment due to their immunoglobulin G structure, but this structure also means most are actively transported across the placenta from the second trimester as fetal passive immunity develops [7–10]. This has led to safety concerns around impacts on the neonate’s immune system. Clinical guidelines have therefore often advised discontinuing treatment by the second trimester where possible [11–15]. The latest updates from the European Alliance of Associations for Rheumatology [16] and the European Crohn’s and Colitis Organization [17] recommended the use of TNF-inhibitor mAbs throughout pregnancy where needed to manage active disease, based on growing reassuring evidence from observational studies [18]. For other mAbs recommendations were more cautious due to sparse data [16]. The decision to discontinue mAb treatment during pregnancy requires women, couples and clinicians to weigh the risks posed by recurrence of active disease, since active autoimmune conditions may cause adverse pregnancy outcomes [19–21], against the often limited evidence on mAbs’ safety profiles [8]. The limited evidence often causes conflicting advice on whether and when to discontinue in pregnancy.

Limited data describe current dispensing patterns during pregnancy for most mAbs. Some studies have described decreases in mAb dispensing during pregnancy [6,22–27], though all literature in this area is limited by modest sample sizes [6,23], and studies have often focussed on only one autoimmune condition [6,24,28] or only on TNF-inhibitors [26]. To our knowledge, no previous studies have described secular trends in mAb prescribing among reproductive age women. We aim to address this gap, and describe dispensing patterns around pregnancy in recent population-wide data from Lombardy. It is important to monitor these patterns to understand current clinical practice, particularly given increasing mAb use and the changing landscape of evidence and clinical guidelines.

We aimed to describe: i) secular trends in mAb dispensing in women of reproductive age and during pregnancy, ii) individual trajectories of mAb dispensation before, during and after pregnancy, iii) prevalence of mAb discontinuation, switching and resumption among pre-pregnancy users, and iv) factors associated with discontinuation of mAbs during pregnancy.

## Methods

### Data source

We used electronic health record data from Lombardy, the largest and most industrialised of Italian regions with a population of approximately 10 million spanning both rural and metropolitan areas. We used an existing cohort of women resident in Lombardy on 01.12.2019 aged 14-49. Data covered the period from 01.01.2012 to 31.12.2024, and included linked demographic information, in-patient hospital admissions, delivery records (covering pregnancies reaching 20 weeks’ gestation), out-patient and emergency room hospital admissions, drugs dispensed in hospitals and reimbursed by the Italian National Health Service, and reasons for exemptions from dispensation payments. Key variables and their data sources are outlined in **Table S1**.

### Study population

We used different analysis subsamples of this cohort for our aims: Aim 1: a) all women of reproductive age (14-49), and then b) all pregnancies. Aim 2: a) all pregnancies, and then b) all pregnancies to women who received a mAb dispensation in the year before pregnancy, during pregnancy, or the year after pregnancy. Aims 3-4: all pregnancies to women who received a mAb dispensation in the period between 1 year before and 90 days before pregnancy, and defined these women as ‘pre-existing users’ for this pregnancy. The subsamples for each aim are illustrated in **Figure S1.**

### Monoclonal antibody dispensing

We previously [21] identified a list of autoimmune conditions which may be diagnosed in women of reproductive age [1]. We searched DrugBank [29] in order to identify mAbs which are used to manage these autoimmune conditions. Full details of the search strategy are outlined in **Supplementary methods** [21,1,29,30] and **Figure S2.**

We identified 23 mAbs (**Table S2**) used to treat seven key autoimmune conditions: inflammatory bowel disease, rheumatoid arthritis, ankylosing spondylitis, psoriasis, multiple sclerosis, systemic lupus erythematosus, and myasthenia gravis. Relationships between indications, mAbs and their targets are summarised in **Figure S3**.

### Exposure periods

For each pregnancy, we used date of delivery and gestational age at birth to estimate the conception date. We then calculated dates for each trimester and approximately 90-day windows in the year before conception and year after delivery [31], which we refer to as ‘periods’ for simplicity. Definitions for these periods are given in **Table S3**. We used the mAb dispensation start date and duration to identify dispensations overlapping with a given period. For details of dispensation data cleaning see **Supplementary methods.**

### Autoimmune conditions

Autoimmune condition diagnosis was derived from any of: 1) outpatient hospital admissions, 2) emergency room admissions, or 3) reasons for payment exemptions. From admissions records we used diagnoses given as either primary reason for admission or secondary diagnoses. ICD-9 codes and exemption reasons used are described in **Table S4.**

### Statistical analyses

We first calculated the prevalence of receiving a mAb dispensation among women of reproductive age in each calendar year, for any mAb and each mAb individually. Since the age distribution of the study cohort changed slightly over the study period (see **Figure S4**), we additionally performed this analysis stratified by 5-year age categories. We then estimated the prevalence of receiving a mAb dispensation during pregnancy for each calendar year of delivery, similarly for any mAb and each individually. For these secular trend analyses, 95% confidence intervals were estimated using the exact method [32].

We additionally described the prevalence of dispensations across periods or trimesters across the year before, during, and the year after pregnancy. We constructed 95% confidence intervals which accounted for multiple pregnancies using robust standard errors calculated by the sandwich R package [33].

To describe trajectories, we assigned each period in each pregnancy to a mAb or none, using the mAb with the longest dispensation duration in cases of multiple exposure.

We then described the prevalence of mAb dispensing patterns around pregnancy, among pre-existing users. We defined dispensing patterns using the pattern of mAb exposure(s) assigned to each period. We defined discontinuation as at least one dispensation in the period between 1 year before and 90 days before pregnancy, followed by any period without mAb exposure between the 90 days pre-pregnancy and the third trimester inclusive. The timing of discontinuation was the first of these periods during which the individual received no mAb dispensation. Switching was defined as changing mAb assigned to a period, for any period between the 90 days pre-pregnancy and the third trimester inclusive. Resumption after pregnancy was defined as discontinuation followed by using any mAb in at least one post-pregnancy period.

To investigate sociodemographic differences in these trends, we repeated analyses stratified by maternal factors related to differential comorbidities and access to health services. These were: maternal age at delivery, country of birth, maternal education as a proxy for socioeconomic position [34], and calendar year (**Table S1**).

We used logistic regression adjusting for calendar year of pregnancy only, to explore the associations of mAbs discontinuation factors including: country of birth (Italy/other), age at delivery, education, employment status, use of assisted reproductive technology, parity (nulliparous/multiparous), previous miscarriage, autoimmune condition, drug dispensed before pregnancy, and European Medicines Agency (EMA) approval year for drug dispensed before pregnancy (**Table S1**). Each model was restricted to complete cases and clustered by pregnant individual to account for multiple pregnancies [33]. We had planned to investigate dispensing centre as a factor but this was not feasible since most dispensations originated from a single centre. We also planned to describe the association of the same factors with switching mAb during pregnancy, but too few women switched mAbs for this analysis to be included.

## Results

We identified 3,049,175 eligible women, who had 858,699 pregnancies between 2012 and 2024 in the Lombardy region (**Table 1**). Mean age at delivery was 32.7 years (standard deviation=5.2). We identified 172,729 mAb dispensations in the study period, with a median dispensation duration of 57 days (interquartile range=30-110 days).

**Table 1.**
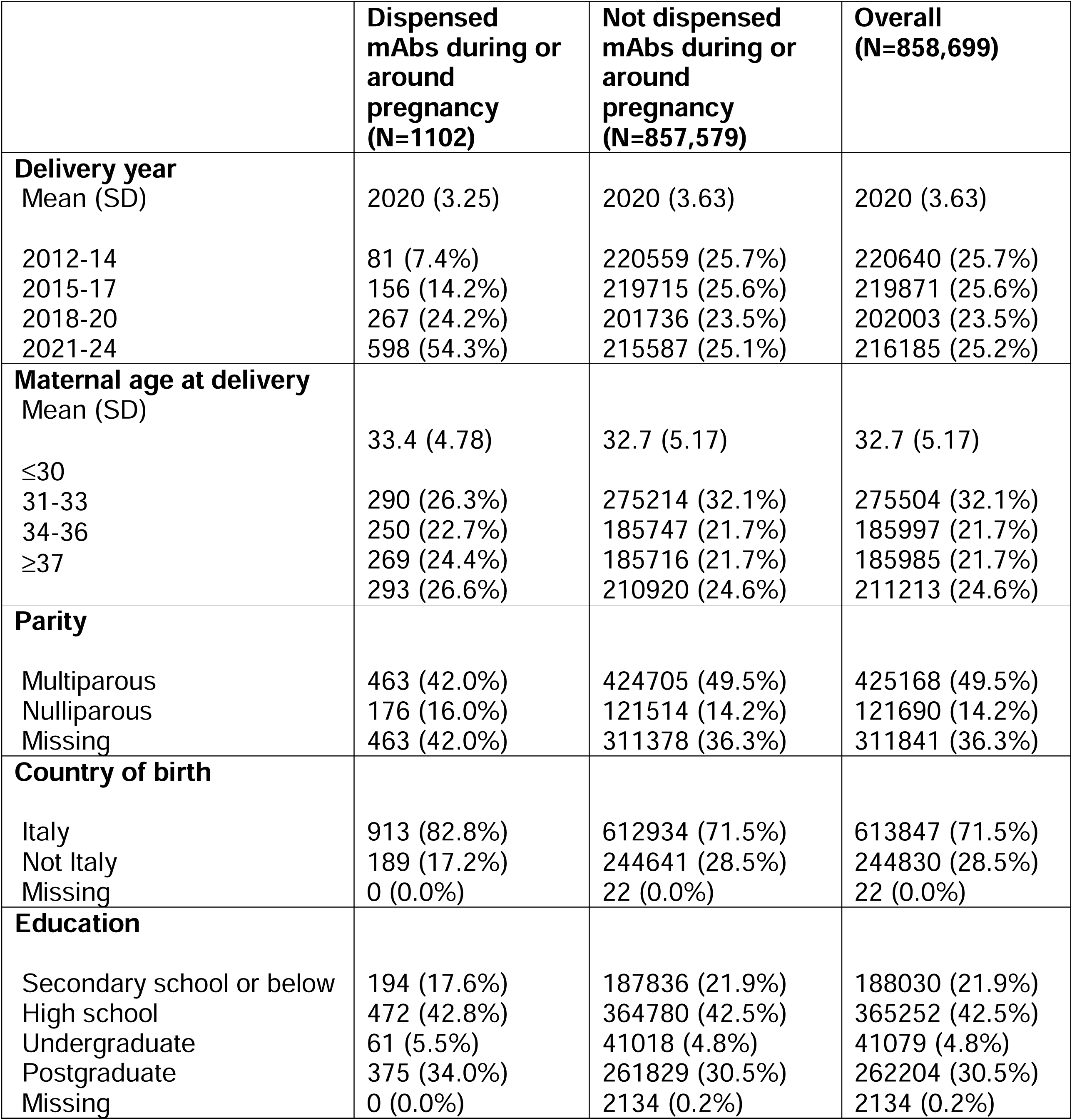
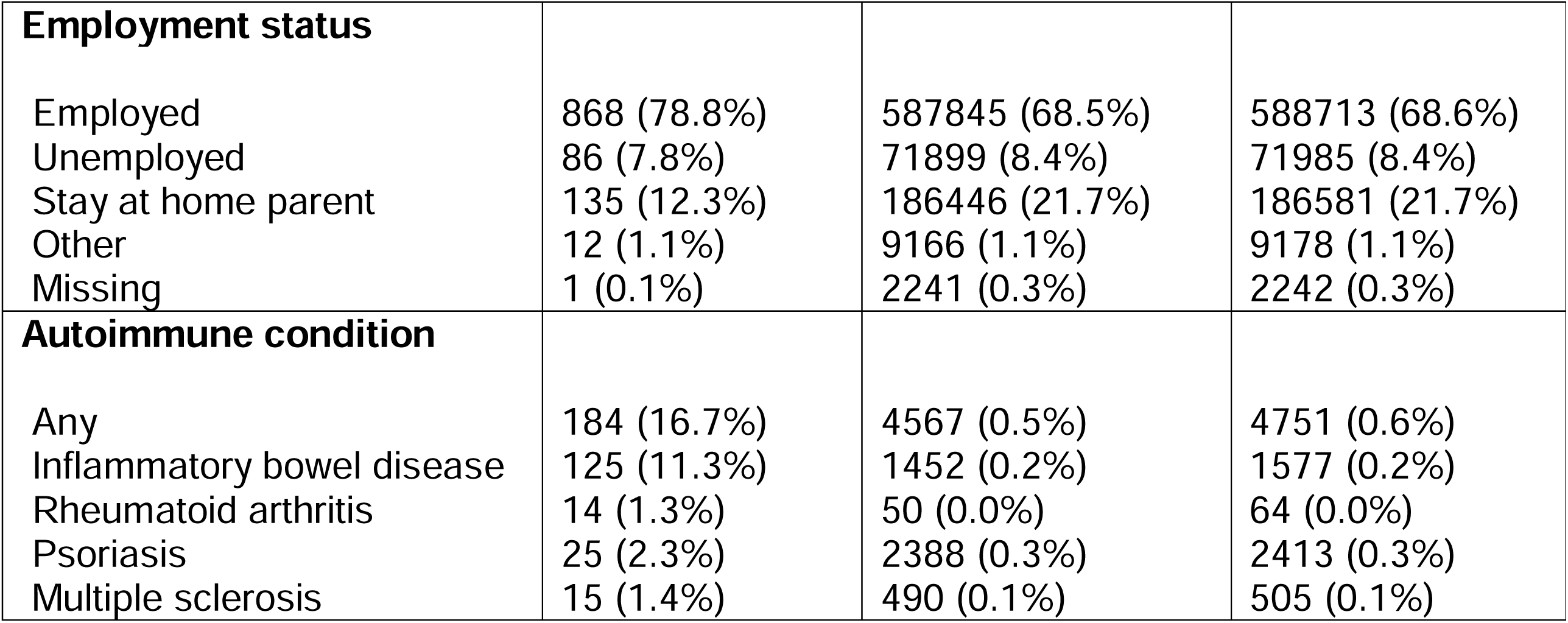
Characteristics of pregnancies in Lombardy cohort in the study period 2012-2024, stratified by those dispensed mAbs during pregnancy, in the year before pregnancy, and/or in the year post-partum (‘Exposed’) and not (‘Unexposed’). Autoimmune condition diagnoses were derived from secondary care and/or dispensation payment exemption records.

|  | **Dispensed mAbs during or around pregnancy (N=1102)** | **Not dispensed mAbs during or around pregnancy (N=857,579)** | **Overall (N=858,699)** |
| --- | --- | --- | --- |
| **Delivery year** |  |  |  |
| Mean (SD) | 2020 (3.25) | 2020 (3.63) | 2020 (3.63) |
| 2012-14 | 81 (7.4%) | 220559 (25.7%) | 220640 (25.7%) |
| 2015-17 | 156 (14.2%) | 219715 (25.6%) | 219871 (25.6%) |
| 2018-20 | 267 (24.2%) | 201736 (23.5%) | 202003 (23.5%) |
| 2021-24 | 598 (54.3%) | 215587 (25.1%) | 216185 (25.2%) |
| **Maternal age at delivery** |  |  |  |
| Mean (SD) | 33.4 (4.78) | 32.7 (5.17) | 32.7 (5.17) |
| ≤30 |  |  |  |
| 31-33 | 290 (26.3%) | 275214 (32.1%) | 275504 (32.1%) |
| 34-36 | 250 (22.7%) | 185747 (21.7%) | 185997 (21.7%) |
| ≥37 | 269 (24.4%) | 185716 (21.7%) | 185985 (21.7%) |
|  | 293 (26.6%) | 210920 (24.6%) | 211213 (24.6%) |
| **Parity** |  |  |  |
| Multiparous | 463 (42.0%) | 424705 (49.5%) | 425168 (49.5%) |
| Nulliparous | 176 (16.0%) | 121514 (14.2%) | 121690 (14.2%) |
| Missing | 463 (42.0%) | 311378 (36.3%) | 311841 (36.3%) |
| **Country of birth** |  |  |  |
| Italy | 913 (82.8%) | 612934 (71.5%) | 613847 (71.5%) |
| Not Italy | 189 (17.2%) | 244641 (28.5%) | 244830 (28.5%) |
| Missing | 0 (0.0%) | 22 (0.0%) | 22 (0.0%) |
| **Education** |  |  |  |
| Secondary school or below | 194 (17.6%) | 187836 (21.9%) | 188030 (21.9%) |
| High school | 472 (42.8%) | 364780 (42.5%) | 365252 (42.5%) |
| Undergraduate | 61 (5.5%) | 41018 (4.8%) | 41079 (4.8%) |
| Postgraduate | 375 (34.0%) | 261829 (30.5%) | 262204 (30.5%) |
| Missing | 0 (0.0%) | 2134 (0.2%) | 2134 (0.2%) |
| **Employment status** |  |  |  |
| Employed | 868 (78.8%) | 587845 (68.5%) | 588713 (68.6%) |
| Unemployed | 86 (7.8%) | 71899 (8.4%) | 71985 (8.4%) |
| Stay at home parent | 135 (12.3%) | 186446 (21.7%) | 186581 (21.7%) |
| Other | 12 (1.1%) | 9166 (1.1%) | 9178 (1.1%) |
| Missing | 1 (0.1%) | 2241 (0.3%) | 2242 (0.3%) |
| **Autoimmune condition** |  |  |  |
| Any | 184 (16.7%) | 4567 (0.5%) | 4751 (0.6%) |
| Inflammatory bowel disease | 125 (11.3%) | 1452 (0.2%) | 1577 (0.2%) |
| Rheumatoid arthritis | 14 (1.3%) | 50 (0.0%) | 64 (0.0%) |
| Psoriasis | 25 (2.3%) | 2388 (0.3%) | 2413 (0.3%) |
| Multiple sclerosis | 15 (1.4%) | 490 (0.1%) | 505 (0.1%) |

Among all pregnancies, 1,102 (0.13%) received a mAb dispensation in the year before pregnancy, during pregnancy, and/or in the year after pregnancy. Pregnancies exposed to mAbs were more likely to be to mothers who had an autoimmune condition diagnosis recorded in secondary care or dispensation payment exemptions (16.7% versus 0.5%), which were largely records of inflammatory bowel disease (11.3% versus 0.2%) (**Table 1**). They were also more likely to be born in 2021-2024 (54.3% versus 25.1%), and to mothers who were born in Italy (82.8% versus 71.5%), had postgraduate-level education (34.0% versus 30.5%), and were employed (78.8% versus 68.5%).

### Secular trends in dispensing

Among women of reproductive age, the prevalence of receiving a mAb dispensation increased from 0.049% (95%CI: 0.045, 0.053) in 2012 to 0.40% (95%CI: 0.39, 0.41) in 2024, consistently across all mAbs (**Figure 1A**) and age groups (**Figures S5** and **S6**). Adalimumab was the most dispensed mAb overall, dispensed to 0.099% (95%CI: 0.094, 0.10) of reproductive age women in 2024.

**Figure 1.**
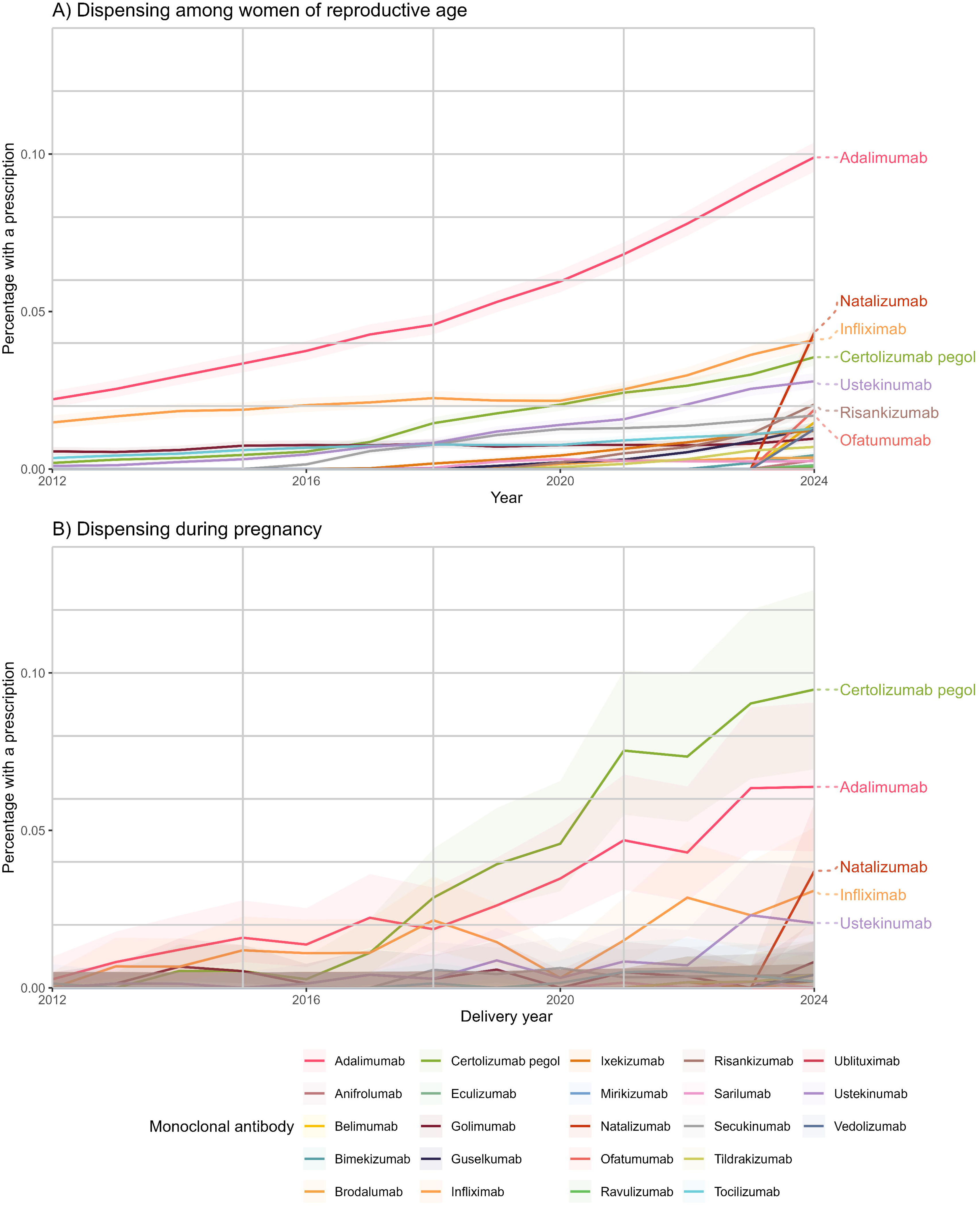
Secular trends in dispensing of monoclonal antibodies in reproductive age women and pregnant women. A) All women of reproductive age by calendar year. B) Women during pregnancy by calendar year of delivery. Drug name refers to active ingredient, which may be administered as different pharmaceutical products.

During pregnancy, there was also an increase in prevalence of mAb dispensing over the study period from 0.0041% (95%CI: 0.00084, 0.012) in 2012 to 0.27% (95%CI: 0.23, 0.32) in 2024. This trend was consistent for the year before and the year after pregnancy (**Figure S7**), and similarly across maternal education and age at delivery, but dispensing was consistently slightly higher among women born in Italy (**Figure S8**). From 2018 onwards certolizumab pegol was the most dispensed mAb during pregnancy (0.095% in 2024; 95%CI: 0.069, 0.13; **Figure 1B**), while adalimumab was the second-most dispensed (0.064% in 2024; 95%CI: 0.043, 0.091). Natalizumab dispensing increased rapidly from 2023 to 0.037% (95%CI: 0.022, 0.059) of pregnancies in 2024.

### Longitudinal patterns of dispensing during and around pregnancy

Prevalence of any mAb dispensation decreased during pregnancy, from 0.080% (95%CI: 0.074, 0.087) before pregnancy to 0.064% (95%CI: 0.058, 0.070) and 0.051% (95%CI: 0.046, 0.057) during the second and third trimesters, respectively (**Figure 2A**). Prevalence also increased slightly after pregnancy, with dispensing in the period one-year pre-pregnancy at 0.071% (95%CI: 0.065, 0.077) compared to 0.090% (95%CI: 0.083, 0.097) one-year postpartum.

**Figure 2.**
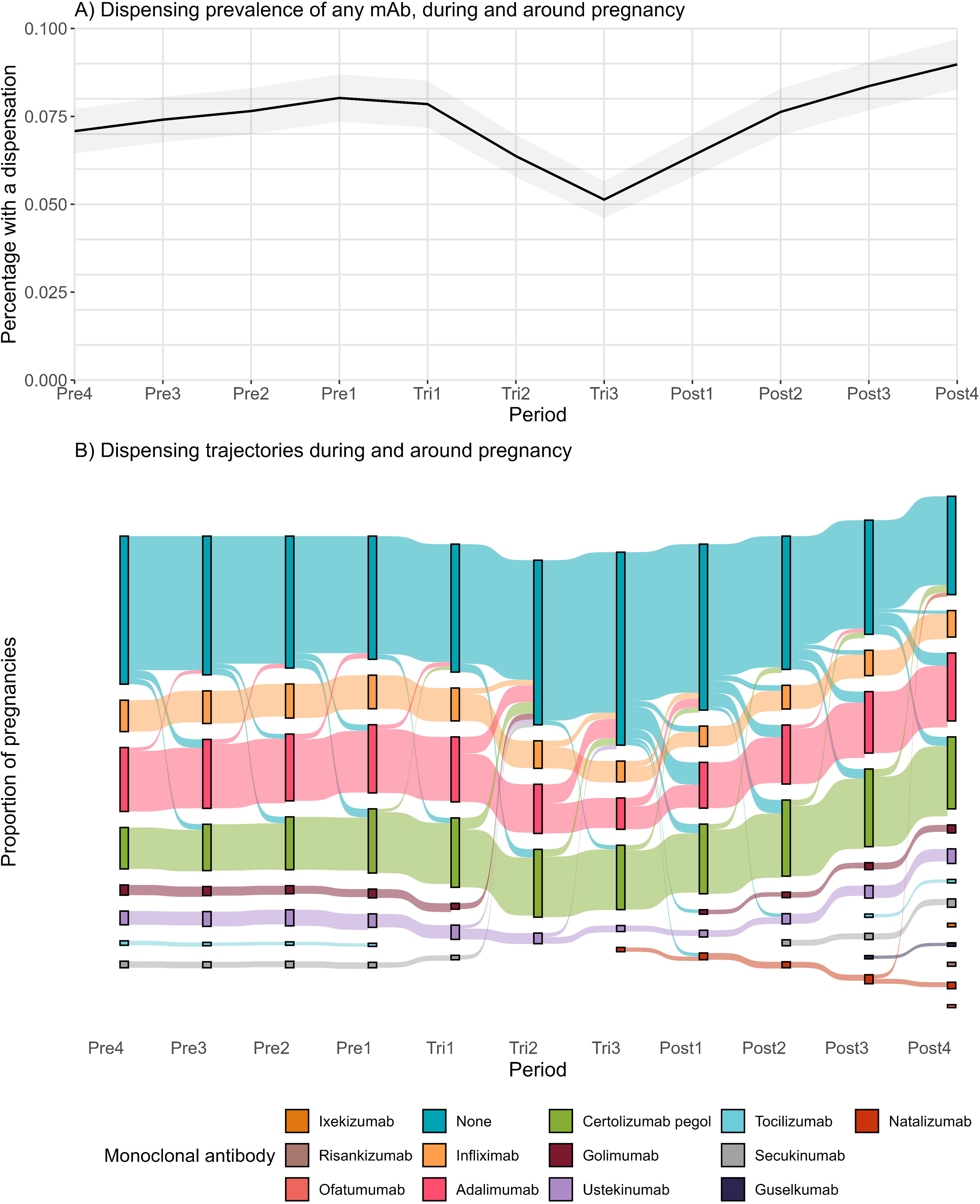
Changes in mAb dispensations before, during and around pregnancy. **A)** Prevalence of any mAb dispensation before, during and after pregnancy for period, with 95% confidence interval accounting for multiple pregnancies to one woman. **B)** Trajectories of mAb dispensations for each trimester or approximately 90-day period in the year before, during and after pregnancy for all pregnant women receiving a mAb in at least one period (n=1,102 pregnancies, n=948 women). The size of the bar at each period is proportional to the count of pregnancies receiving that mAb or none, for all uncensored pregnancies. For periods with multiple mAb dispensations the dispensation with the longest duration in that period was selected (i.e. if someone received adalimumab for 8 days and certolizumab pegol for 27 in one period, the period would be designated as exposed to certolizumab pegol). Some monoclonal antibody drugs are not shown for some periods, because trajectories containing fewer than 10 pregnancies have been censored.

These trends were similar across maternal age at delivery and education (**Figure S9**). For both more recent delivery years and women born in Italy, overall dispensing prevalence during pregnancy was consistently higher and the decrease in the third trimester more pronounced. The absolute decrease in dispensing was greatest for adalimumab and less pronounced for certolizumab pegol and other mAbs (**Figure S10**).

**Figure 2B** illustrates the trajectories of all pregnant women who were exposed to a mAb in one of these periods (n=1,102 pregnancies to 948 women), which similarly found pregnancies were least likely to receive a dispensation overlapping the second (51.1%) and third trimesters (40.9%). For most mAbs, the number of pregnancies exposed decreased between pre-pregnancy and the third trimester, for instance infliximab users (9.7% to 6.4%) and adalimumab users (18.5% to 9.0%). On the other hand, the count of pregnancies exposed to certolizumab pegol remained similar, increasing slightly from 19.1% to 19.4%.

### Prevalence of discontinuation and switching during pregnancy and resumption after pregnancy

Among women who were exposed to a mAb between 1 year before and 90 days before pregnancy (n=722, ‘pre-existing users’), the majority discontinued either before or during pregnancy (n=385/722, 53.3%), many continued to receive mAb dispensations during pregnancy (n=337/722, 46.7%), and a small number switched mAb during pregnancy (n=24/722, 3.3%). Of those who discontinued, just over half resumed any mAb treatment in the year post-delivery (n=215/385, 55.8%).

The most common timing for discontinuation was the second trimester (n=145/385, 37.7%), followed by the third trimester (n=106/385, 27.5%), before pregnancy (n=89/385, 23.1%), and first trimester (n=45/385, 11.7%). **Figure 3** summarises overall dispensing patterns (see **Table S6** for counts), which were largely consistent across strata of maternal factors.

**Figure 3.**
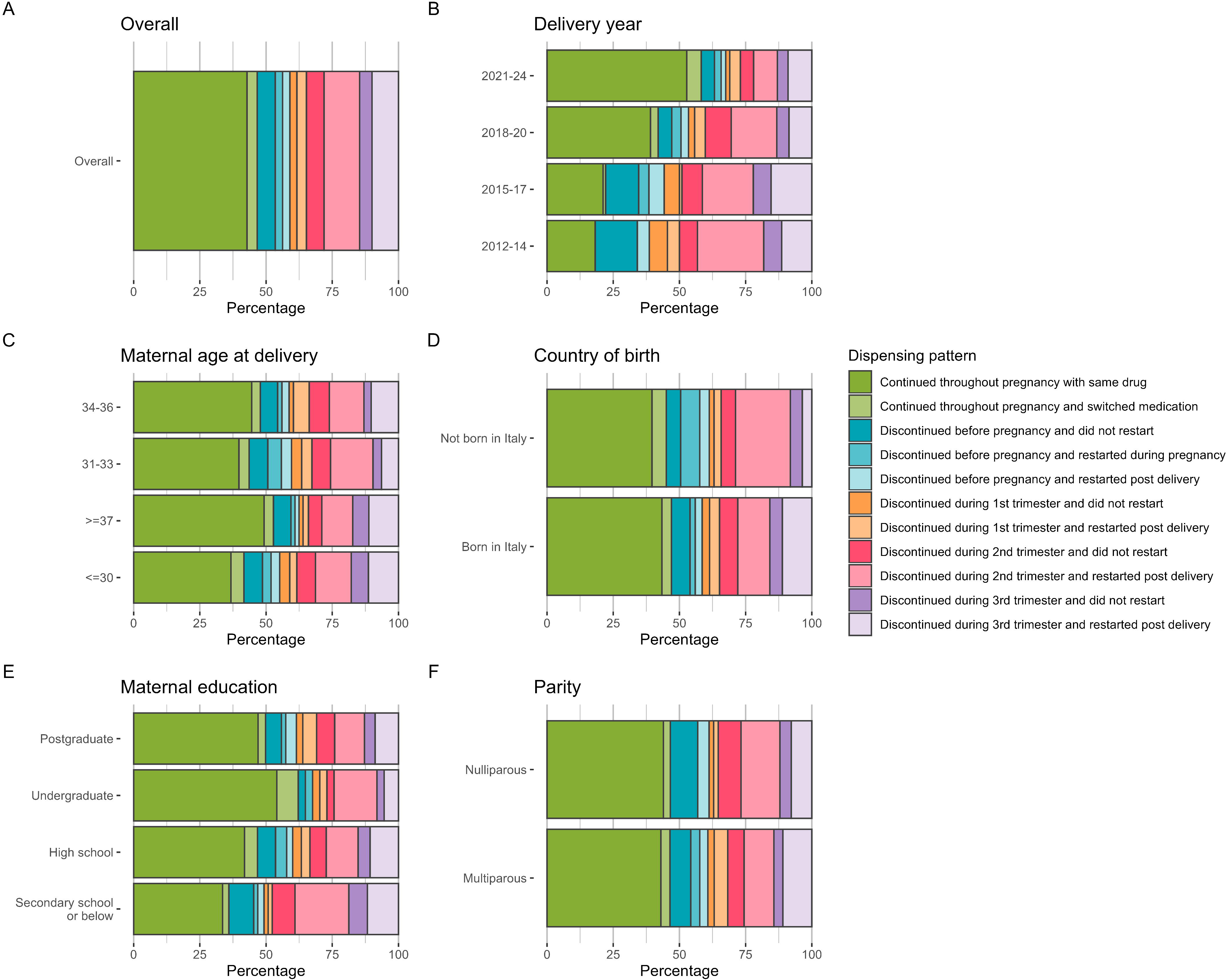
Prevalence of different dispensing patterns before and during pregnancy, among pre-existing users (n=722 pregnancies to 630 women), overall (A) and stratified by maternal factors (B-F). Pre-existing uses were defined as exposed to a mAb between 1 year before and 90 days before pregnancy. A small number of pregnancies which discontinued also switched mAb (n<10). Pregnancies with missing information on variables were discarded, resulting in a reduced sample size for parity (n=409 pregnancies) only.

However, continuation of mAb dispensations throughout pregnancy was more prevalent in later delivery years, increasing from 18.2% (n=8/44) in 2012-14 to 58.3% (233/400) in 2021-2024. There were also expected differences in dispensing patterns dependent on the drug dispensed in the year before pregnancy (**Figure S11**), with certolizumab pegol and multiple mAb users the most likely to continue the same drug throughout all three trimesters (65.3% (113/173) and 50.8% (31/61), respectively).

### Factors associated with discontinuation

Whether the mother was born in Italy, maternal age, education, employment or parity did not appear to influence odds of discontinuation (**Figure S12**). We found evidence that compared to those who had conceived spontaneously, those who had conceived by assisted reproductive technology were more likely to discontinue (OR=1.92, 95%CI:0.98-3.77). We also found evidence that both previous miscarriage (OR=0.68; 95%CI:0.44-1.06) and a recorded autoimmune condition (OR=0.62, 95%CI:0.41-0.96) may be associated with decreased odds of discontinuation.

Compared to pre-existing users dispensed adalimumab in the year before pregnancy, women dispensed infliximab (OR=0.65, 95%CI:0.41-1.04), certolizumab pegol (OR=0.51, 95%CI:0.33-0.81), or multiple drugs (OR=0.35, 95%CI:0.18-0.69) were less likely to discontinue. On the other hand, users of secukinumab (OR=2.57, 95%CI:1.04-6.35), ustekinumab (OR=1.89, 95%CI:1.01-3.54) and other mAbs (OR=4.35, 95%CI:1.86-10.19) were more likely to discontinue. We found evidence that compared to mAbs approved by the European Medicines Agency in 1999 (infliximab), those dispensed mAbs approved in 2003 (adalimumab) and 2015 (secukinumab) were more likely to discontinue (2003 OR=1.55, 95%CI:0.97-2.49; 2015 OR=4.05, 95%CI:1.56-10.51), while there was more modest evidence for an association for those approved in 2009 (certolizumab pegol, ustekinumab, golimumab; OR=1.27, 95%CI:0.80-2.04).

## Discussion

### Main findings

We identified substantial increases in the prevalence of mAb dispensing among both women of reproductive age and pregnant women, between 2012-2024 in Lombardy. In 2024, certolizumab pegol was the most widely used mAb during pregnancy, followed by adalimumab and natalizumab. For individual pregnancies, there was a decrease in dispensing from before pregnancy to the third trimester, which recovered post-partum. Among pre-existing users, we found one in thirty switched mAbs and around half discontinued before or during pregnancy; among those who discontinued, approximately half went on to receive a mAb dispensation in the year after pregnancy.

The prevalence of discontinuation decreased over time, and varied between mAbs. Discontinuation was least common for women who had used certolizumab pegol or multiple drugs before pregnancy. We found modest evidence that discontinuation was more likely for pregnancies conceived using assisted reproductive technology, and negatively associated with a recorded autoimmune condition diagnosis or previous miscarriage, but no evidence for associations with other health or socioeconomic factors.

### Interpretation

Few previous studies have estimated the prevalence of mAb use in population-wide data, and as far as we are aware none have estimated this in women of reproductive age or during pregnancy. Our estimates for the prevalence of mAb dispensations were compatible with previous population-wide studies of prevalence for both autoimmune conditions [1] and mAbs [5]. In accordance with our results, a French study of multiple sclerosis management during pregnancy similarly found increased use of natalizumab in recent years [4]. The increase in mAb dispensations may reflect increases in autoimmune condition diagnoses [1,35], and/or rising use and availability of mAbs [3–5,26].

We found that mAbs were discontinued in around half of pregnancies, in accordance with previous studies across Europe and North America [6,22–27]. In line with our findings, a previous Italian study found that mAb use declined during pregnancy across subspecialties [22]. Another study of Italian women in the Lazio region found that women with psoriasis all discontinued mAb treatment during pregnancy, as did the majority of women with rheumatoid arthritis [36]. Their findings may differ from ours due to their smaller sample size (<100 women per condition using mAbs), focus on psoriasis treatments, older dataset (until 2016), and/or regional differences in healthcare provision [22].

We found a modest increase in the prevalence of mAb dispensation between before and after pregnancy; however, a previous study found that prevalence of mAb use in the year post-partum was lower than in the year pre-pregnancy [22]. In the present study, we also found that just over half of women who discontinued mAbs during pregnancy later resumed mAb treatment post-partum. The increase in mAb dispensation we saw post-pregnancy is likely to reflect secular trends in mAb use, since we found that mAb use increased rapidly between 2021-2024 and the previous study period ended in 2021 [22]. It might also be explained by post-pregnancy flare-ups in disease activity or post-pregnancy disease onset commonly seen in rheumatic arthritis and multiple sclerosis [37,38].

Across strata of maternal age, country of birth and education, we found that trends were broadly consistent for both secular trends of increased mAb use over time (aim 1) and longitudinal patterns of dispensation during pregnancy (aim 2). mAb use was higher for women born in Italy than abroad, which could reflect differences in age, autoimmune condition prevalence [39], or differential access to healthcare in Lombardy.

Our finding that discontinuation was common in the second and third trimesters, and varied between drugs, aligns with European and Italian guidelines [14–17]. These have advised discontinuing most mAbs at this stage due to the risk of placental antibody transfer, unless the benefits of managed disease outweigh the risk of the drug. Certolizumab pegol use pre-pregnancy was associated with reduced risk of discontinuation. This is in-keeping with guidelines which recommend certolizumab pegol use during pregnancy [16,17] because the molecule does not contain the Fc portion required for active transport across the placenta, though it may still cross via passive diffusion [10,40].

Switching between mAbs was uncommon over the period three months before pregnancy until delivery. As far as we are aware, no prior studies have described switching between mAb treatments during pregnancy. Our findings may reflect minimal switching, or that switching usually occurs earlier than this for planned pregnancies.

mAb discontinuation during pregnancy varied between drugs and decreased between 2012-2024. This change has been reported previously for Italy in data up to 2021 [22] and reflects updates in guidelines [11–13,16] and emerging safety data [18,41]. Safety data is most reassuring for TNF inhibitors [18], and is emerging for natalizumab [42,43], vedolizumab and ustekinumab [41]. On the other hand, it is still lacking for some more novel and less widely used mAbs [8].

We found modest evidence that discontinuation was more likely for pregnancies conceived by assisted reproductive technology and less likely for women with autoimmune diagnoses and previous miscarriage, but unrelated to age or other sociodemographic factors. Similarly, other studies have identified few maternal factors related to mAb discontinuation except drug or calendar year [22,24,44]. Our finding for miscarriage is concordant with a Canadian study which found prior adverse birth outcomes were associated with lower odds of mAb discontinuation in the first trimester (OR=0.22, 95%CI:0.05-0.95) [24].

### Strengths and limitations

This is the first study to describe trends in mAb dispensations among all women of reproductive age and all pregnancies, and to describe switching between mAbs during pregnancy, as far as the authors are aware. We were able to integrate data on secondary care dispensing and delivery records for a cohort of over 3 million women of reproductive age, through to 2024. We are confident that we captured the vast majority of mAbs dispensed in Lombardy since we have access to complete data of hospital-based drug dispensing, which is the standard approved practice for dispensation of this class of drugs in Italy.

A limitation was the modest sample size of women dispensed mAbs, since mAbs are reserved to treat moderate to severe autoimmune conditions. We did not have complete data on autoimmune condition diagnoses, since primary care records were not available. However, the vast majority of mAbs studied here are only indicated for autoimmune conditions. The only exceptions were ofatumumab (licensed for chronic lymphocytic leukaemia by the EMA 2010-2019) and tocilizumab (adjunct therapy for Covid-19, and cytokine release syndrome), which only 15 and 30 pregnancies were exposed to throughout the study period, respectively. Importantly, we also lacked data on condition severity, the main factor influencing discontinuation as emphasised in clinical guidelines [16,17]. Moreover, dispensing data may not correspond to medication use in practice. Finally, we could only include pregnancies reaching 20 weeks gestation therefore excluding early pregnancy losses. The findings in this study reflect use of mAbs for autoimmune conditions in Italy and may not generalise to recent trends in other countries, highlighting the importance of replication in different settings.

## Conclusion

We found increasing mAb use during pregnancy over the past decade in Lombardy, Italy. Over time, mAbs were less likely to be discontinued during pregnancy, in line with shifting guidelines and emerging safety data. Further work is needed to understand the impact of increased mAb use on both maternal and fetal outcomes, particularly for novel and rarely-used mAbs.

## Data Availability

Access to the Lombardy region data can be obtained by submitting a project application for individual-level data. The application includes information on the purpose of data use; the requested data, including the variables, definitions of the target and control groups; the dates of the data needed; and a data utilization plan. The requests are evaluated case by case. Once approved, the data are sent to a secure computing environment (Daas 2.0).

## Acknowledgements

This study was carried out according to the Lombardy Region laws on the use of regional healthcare databases for research activities (D.g.r. X/4893, 09/03/2016; D.g.r. XI/491, 02/08/2018; D.g.r. XI/6387, 16/05/2022.), and in particular on COVID-19 disease (D.g.r XI/3019, 30/03/2020). We thank the Struttura Controllo e monitoraggio dati, LEA e outcome and the Azienda Regionale per l’Innovazione e gli Acquisti (ARIA) S.p.a. for the support of this research. The authors would also like to thank the data managers at Lombardy Region and Human Technopole for their helpful assistance. We are grateful to Cinzia Rotondo, Laura Ghirardi, Albert Navarro Gallinad, Lucia Piubeni, and Andrea Corbetta for their technical support and valuable feedback on earlier drafts of this manuscript.

## Contribution to Authorship

EA, MCB, DAL, and LZ conceived and designed the study. EA wrote an initial analysis plan, undertook data analyses and wrote the first draft of the manuscript. LZ established the cohort and was responsible for data oversight and management. EA, VN, RC, BGF, CB, KB, MCB, DAL, and LZ provided comments and critical revisions on the analysis plan and drafts of the manuscript. All authors read and approved the final manuscript.

## Details of Ethics approval

The authors have nothing to report.

