## Supplement for "Monoclonal antibody dispensing during and around pregnancy: a descriptive analysis using electronic health records in Italy"

Supplementary Material

### Methods

#### Monoclonal antibody selection

We have previously [29] identified a list of autoimmune conditions which may be diagnosed in women of reproductive age (defined as those aged 14-49 years), on the basis of a study describing sex-stratified autoimmune condition prevalence [1].

We selected mAbs indicated to manage these autoimmune conditions. First, we mapped mAbs listed in the WHO’s Anatomical Therapeutic Chemical classification ‘L04: Immunosuppressants’ to their indications in adults using DrugBank [29] and the European Medicines Agency (EMA) [30]. We then selected only mAbs used to manage autoimmune conditions which can be diagnosed in women of reproductive age.

We additionally required mAbs to be approved for use in the EMA, and recommended for routine courses of treatment more frequently than every 6 months so that we could accurately capture whether dispensing changed during and around pregnancy.

#### Dispensation data cleaning

Throughout all analyses, we required a mAb dispensation to overlap with a period to define it as exposed. Among all mAb dispensations in the study period, a small proportion (1.7%) of dispensation durations were longer than 365 days and these were trimmed to 365 days to remove implausible data, since mAb injections may be stored and administered at home but are unlikely to last longer than one year.

### Figures

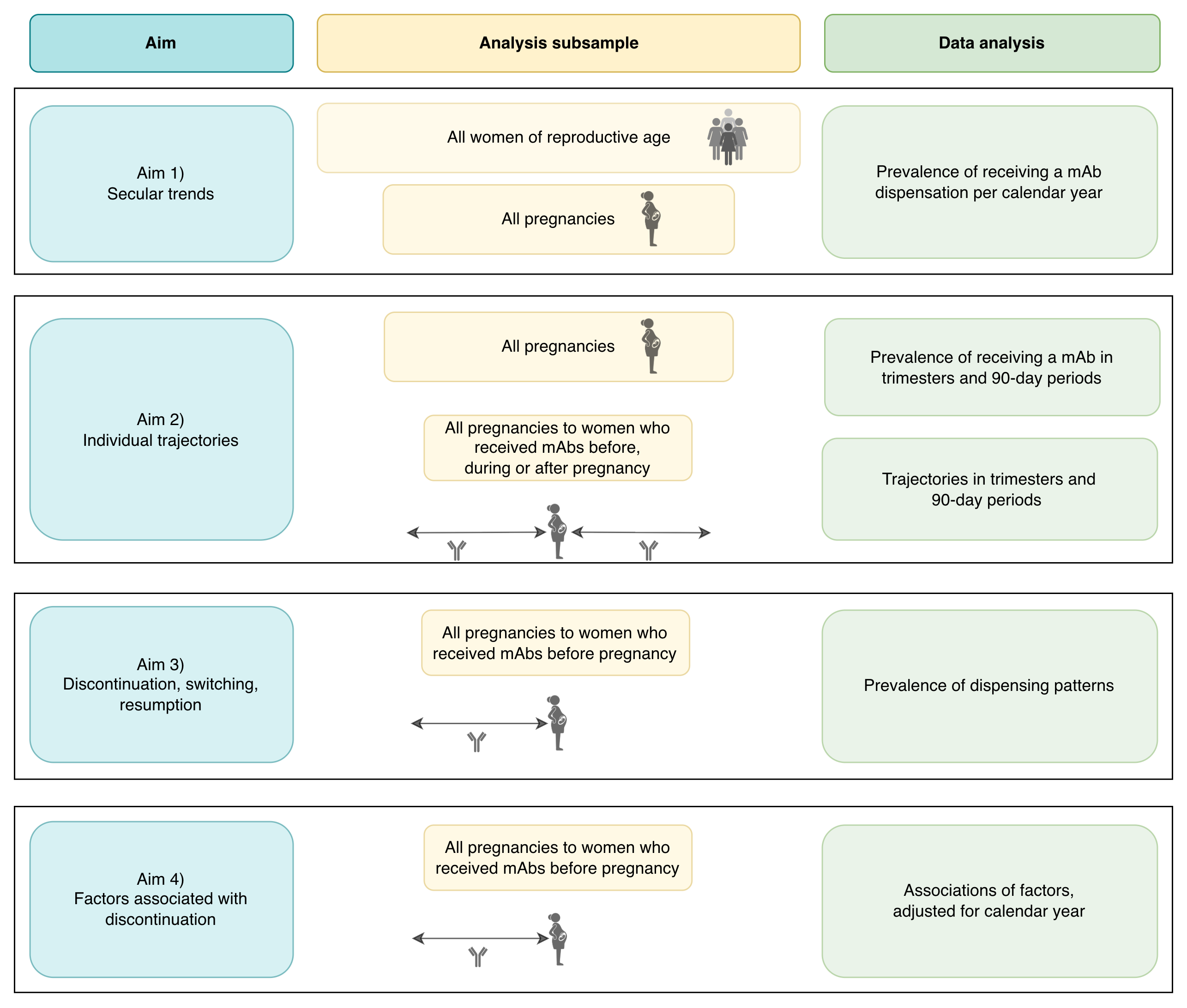

#### Figure S1. Schematic overview of study aims, analysis subsamples, and data analysis.

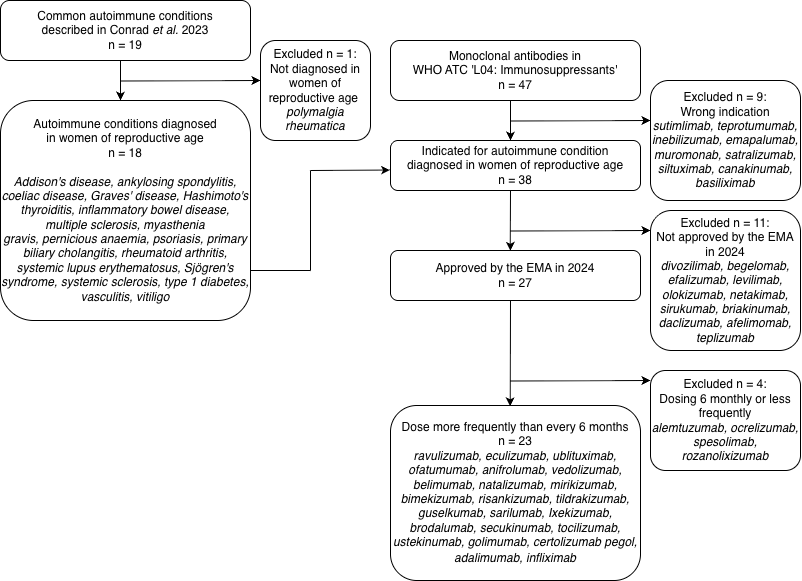

#### Figure S2. Selection of monoclonal antibodies used to manage autoimmune conditions occurring at reproductive age.

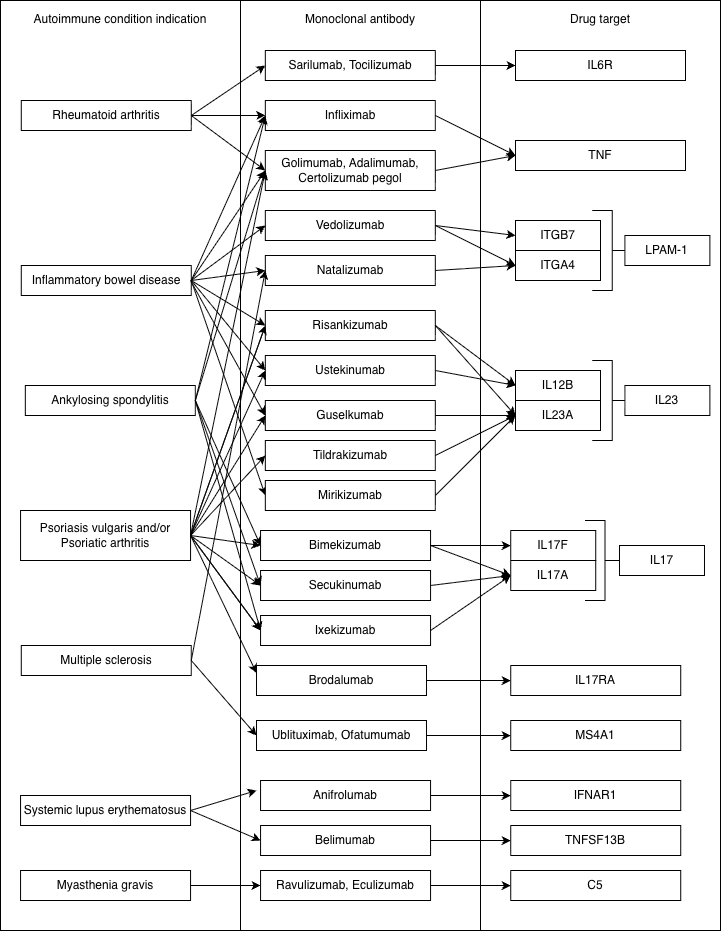

#### Figure S3. Relationships between autoimmune condition indications, monoclonal antibodies of interest, and their targets.

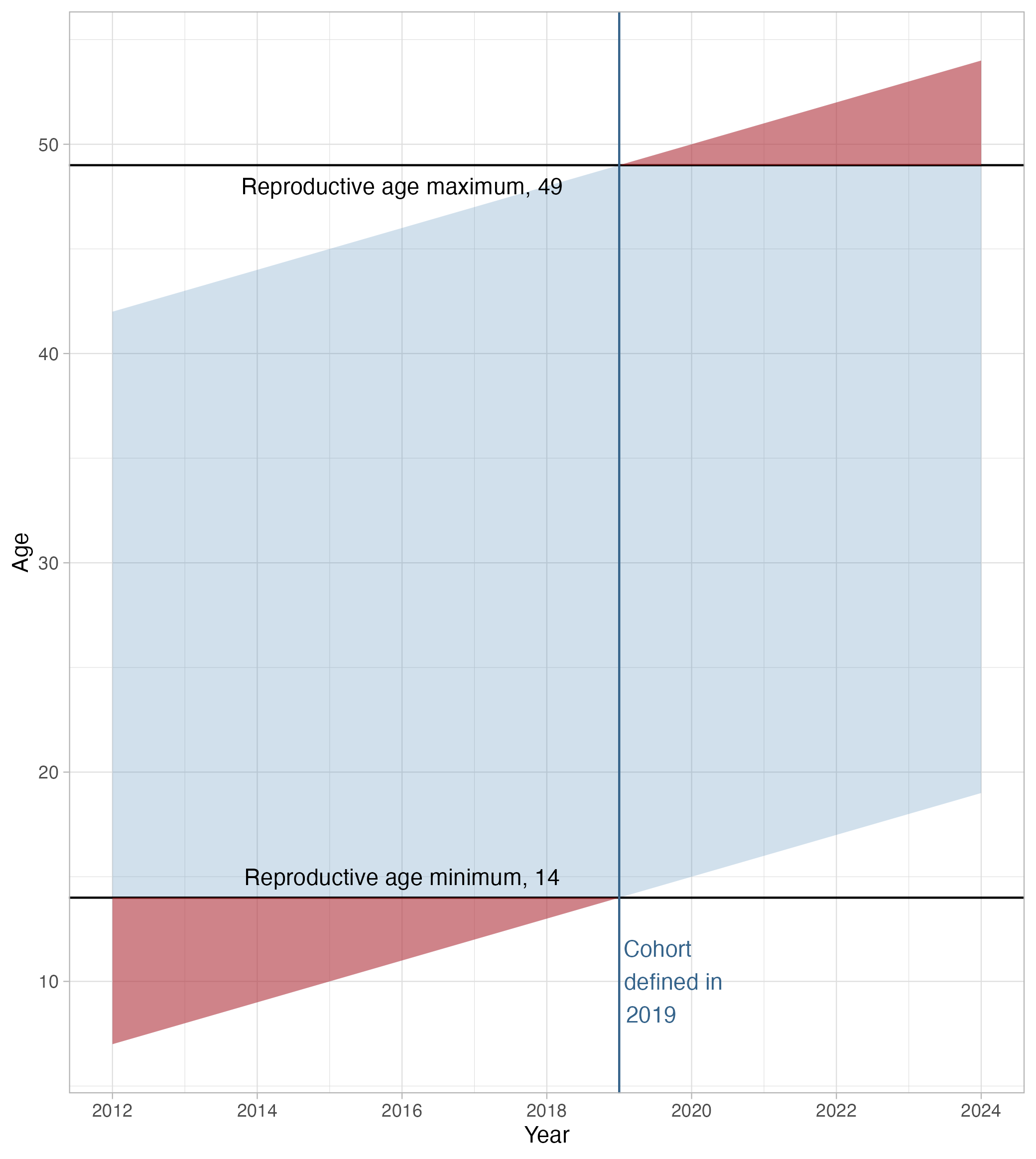

#### Figure S4. Cohort definition and age profile.

Since the cohort was defined as all women of reproductive age resident in Lombardy in 2019, the age profile of the cohort changed slightly over the study period (2012-2024).

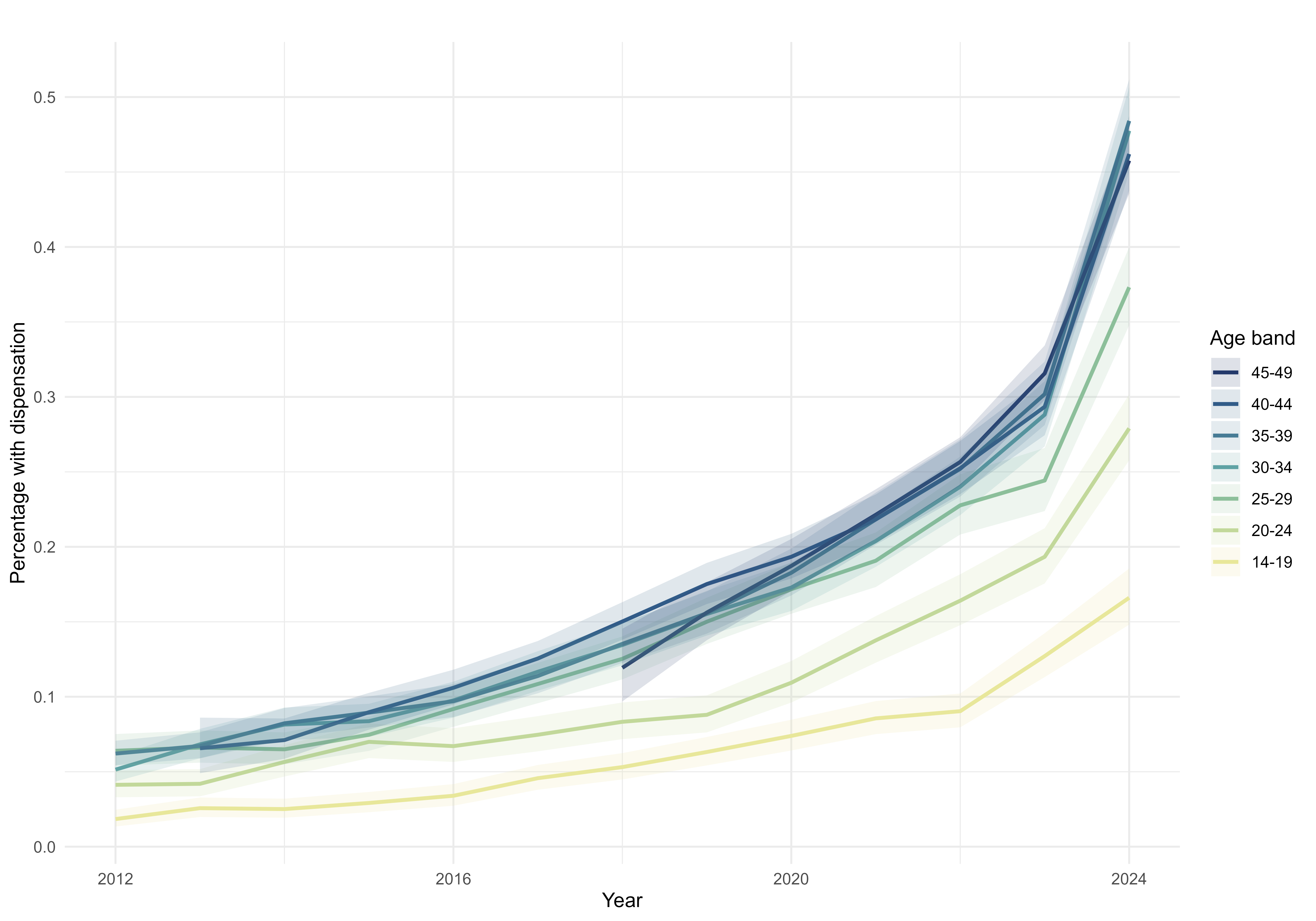

#### Figure S5. Prescribing prevalence of any mAb in each calendar year among women of reproductive age, stratified by age band.

95% confidence intervals shown around each age band. This analysis was performed since the cohort was defined as all women of reproductive age resident in Lombardy in 2019, so the age profile of the cohort changed slightly over the study period.

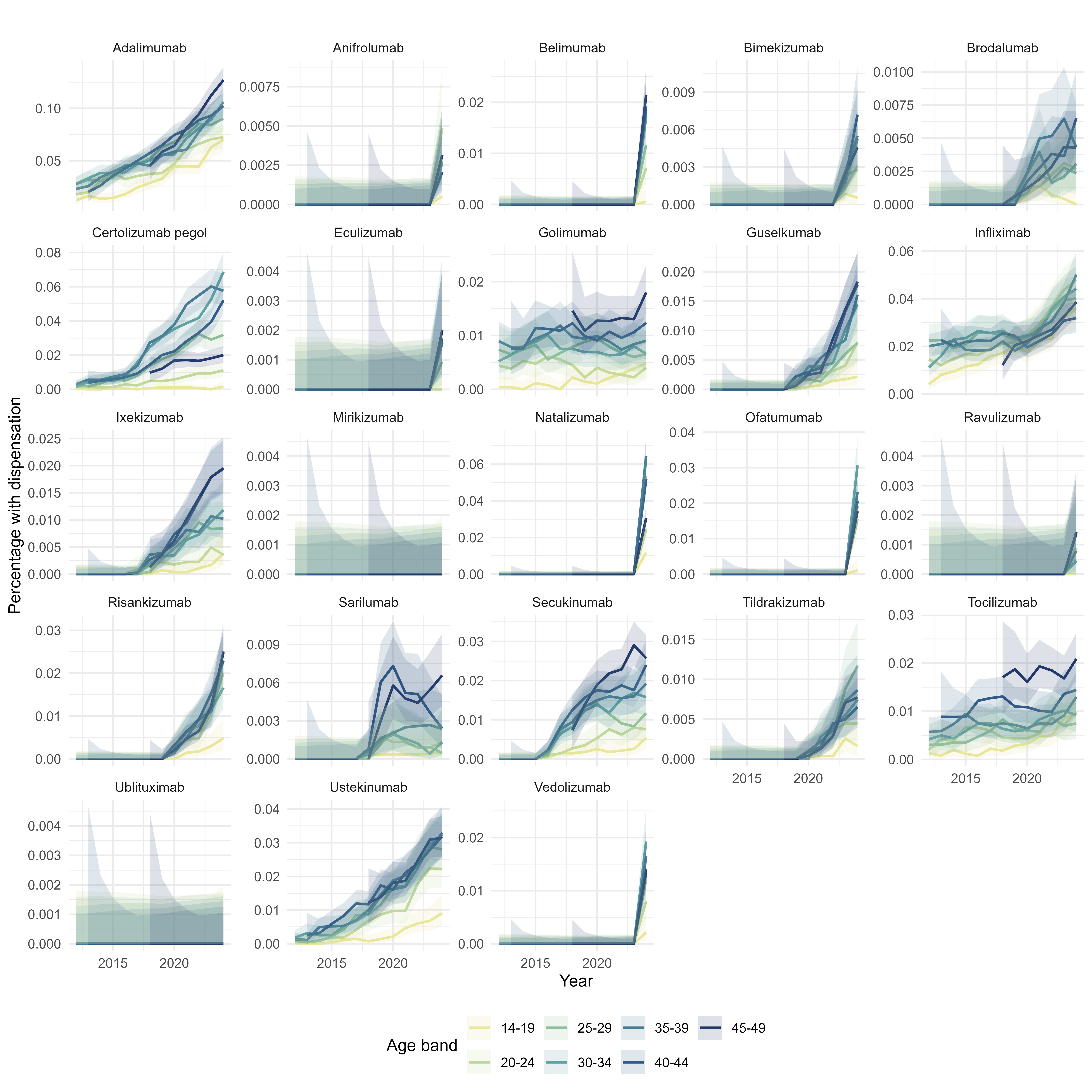

#### Figure S6. Prescribing prevalence of each mAb in each calendar year among women of reproductive age, stratified by age band.

95% confidence intervals shown around each age band. This analysis was performed since the cohort was defined as all women of reproductive age resident in Lombardy in 2019, so the age profile of the cohort changed slightly over the study period.

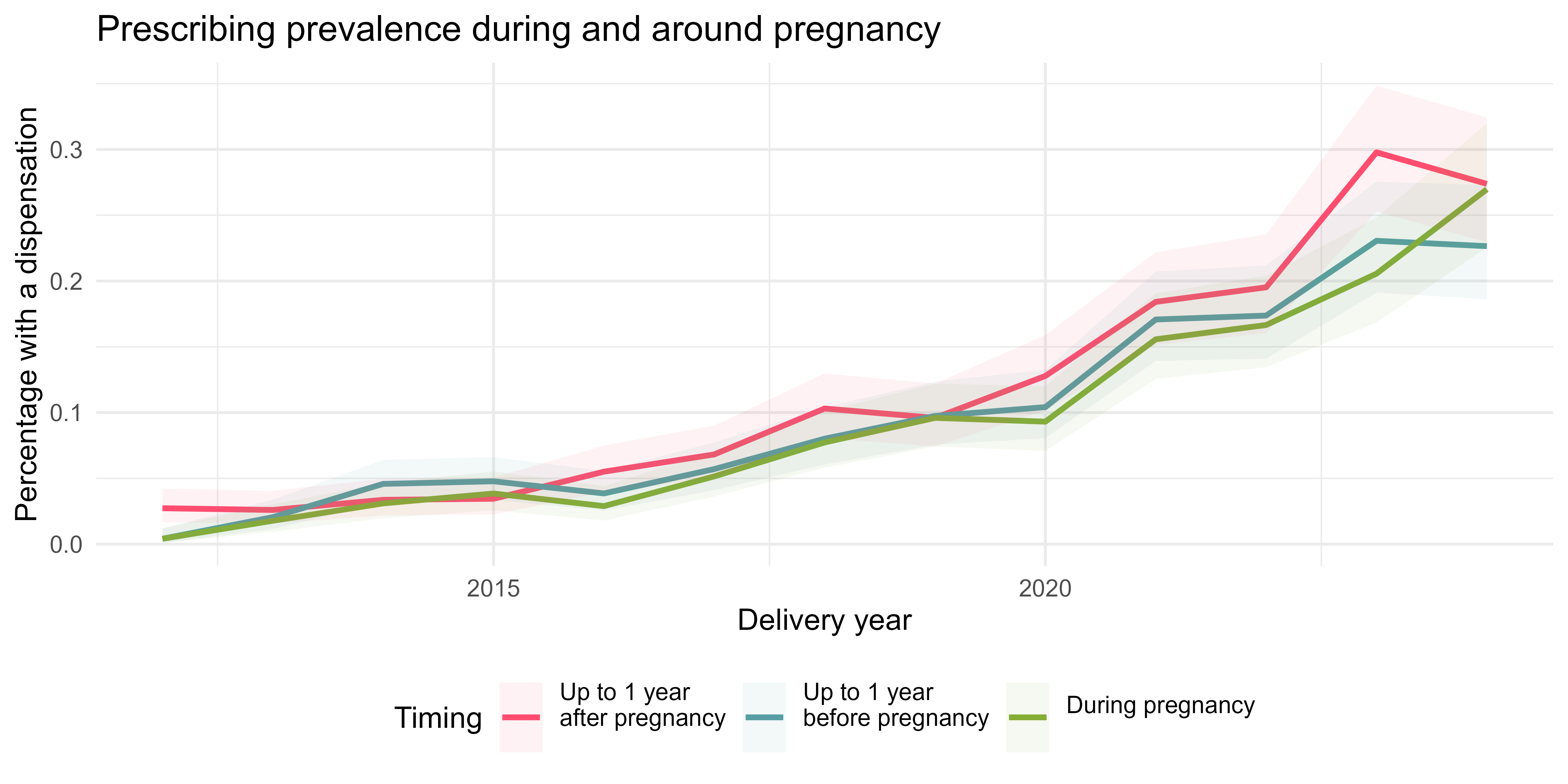

#### Figure S7. Secular trends in prescribing of monoclonal antibodies during and around pregnancy, stratified by period of exposure.

Categories are mutually inclusive, i.e. one pregnancy could be classed as dispensed monoclonal antibodies in the year prior, during and/or after pregnancy. Bands illustrate 95% confidence intervals around annual prevalence estimates.

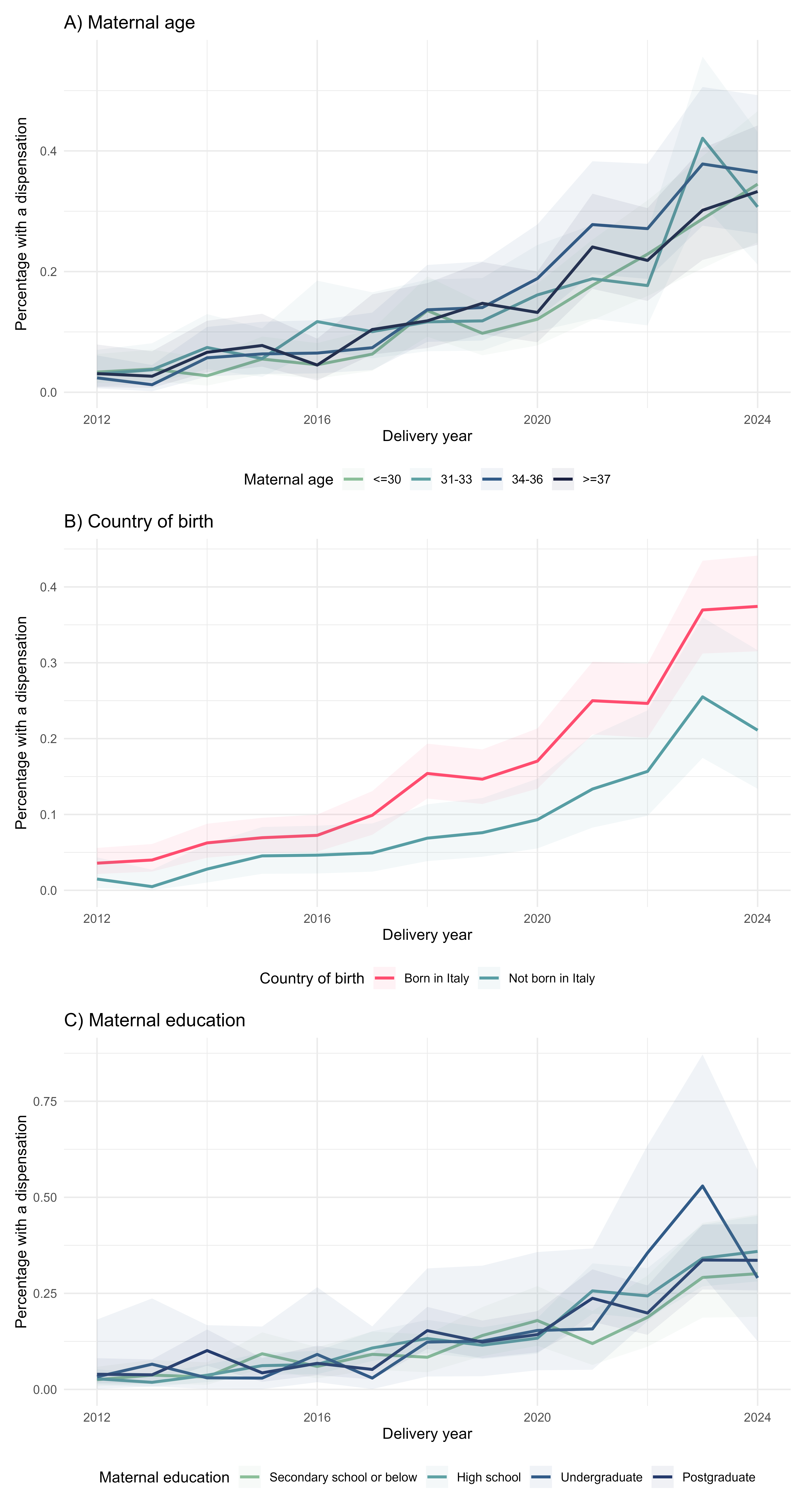

#### Figure S8. Secular trends in prescribing of monoclonal antibodies during pregnancy over time, stratified by maternal characteristics.

A) Maternal age, B) Maternal country of birth, C) Maternal education level. Bands illustrate 95% confidence intervals around annual prevalence estimates.

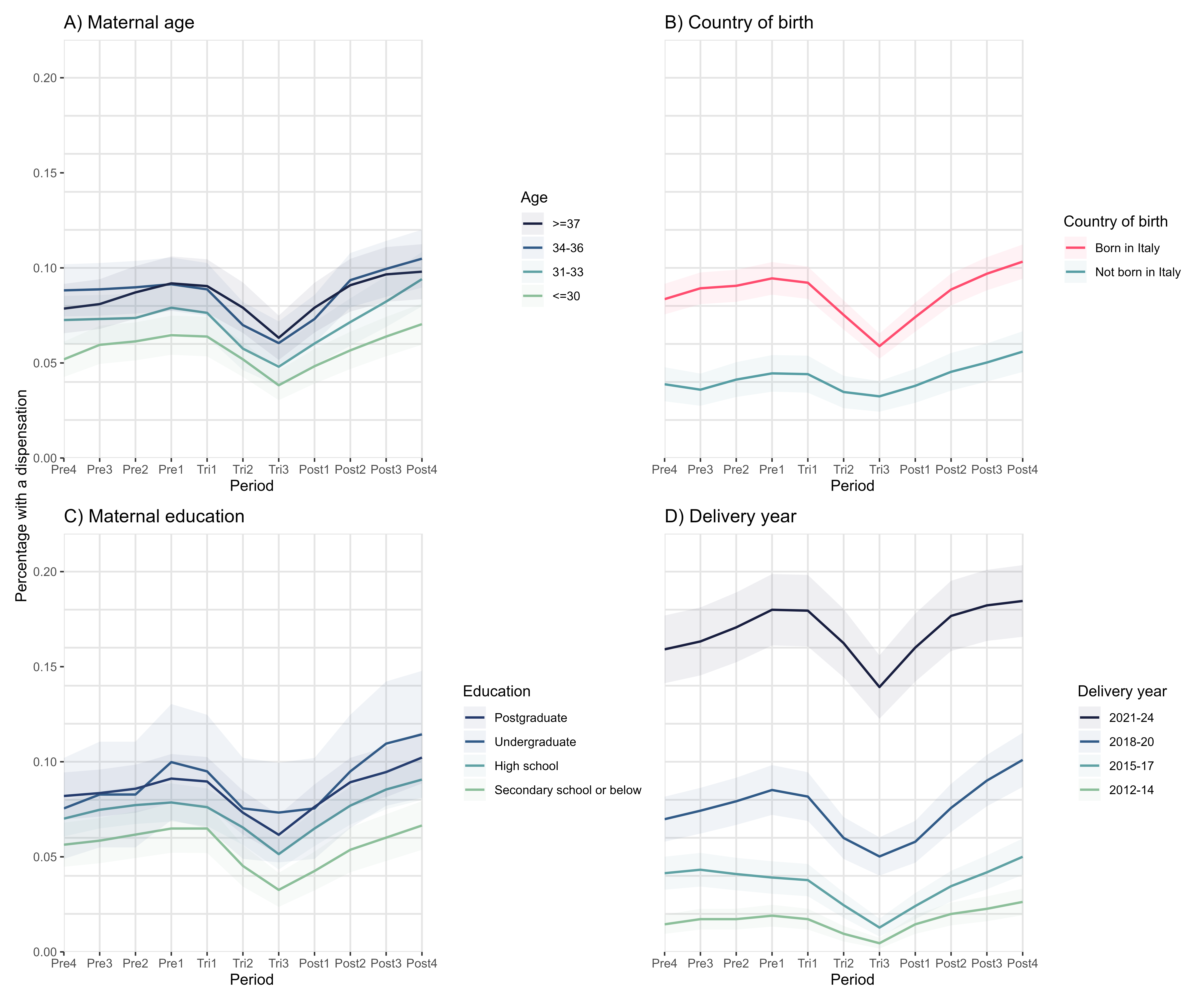

#### Figure S9. Prevalence of monoclonal antibody prescribing during and around pregnancy by period, stratified by maternal characteristics.

A) Maternal age, B) Maternal country of birth, C) Maternal education level. Bands illustrate 95% confidence intervals around annual prevalence estimates, accounting for multiple pregnancies to the same individual. Denominator is all pregnancies in the study period (2012-2024).

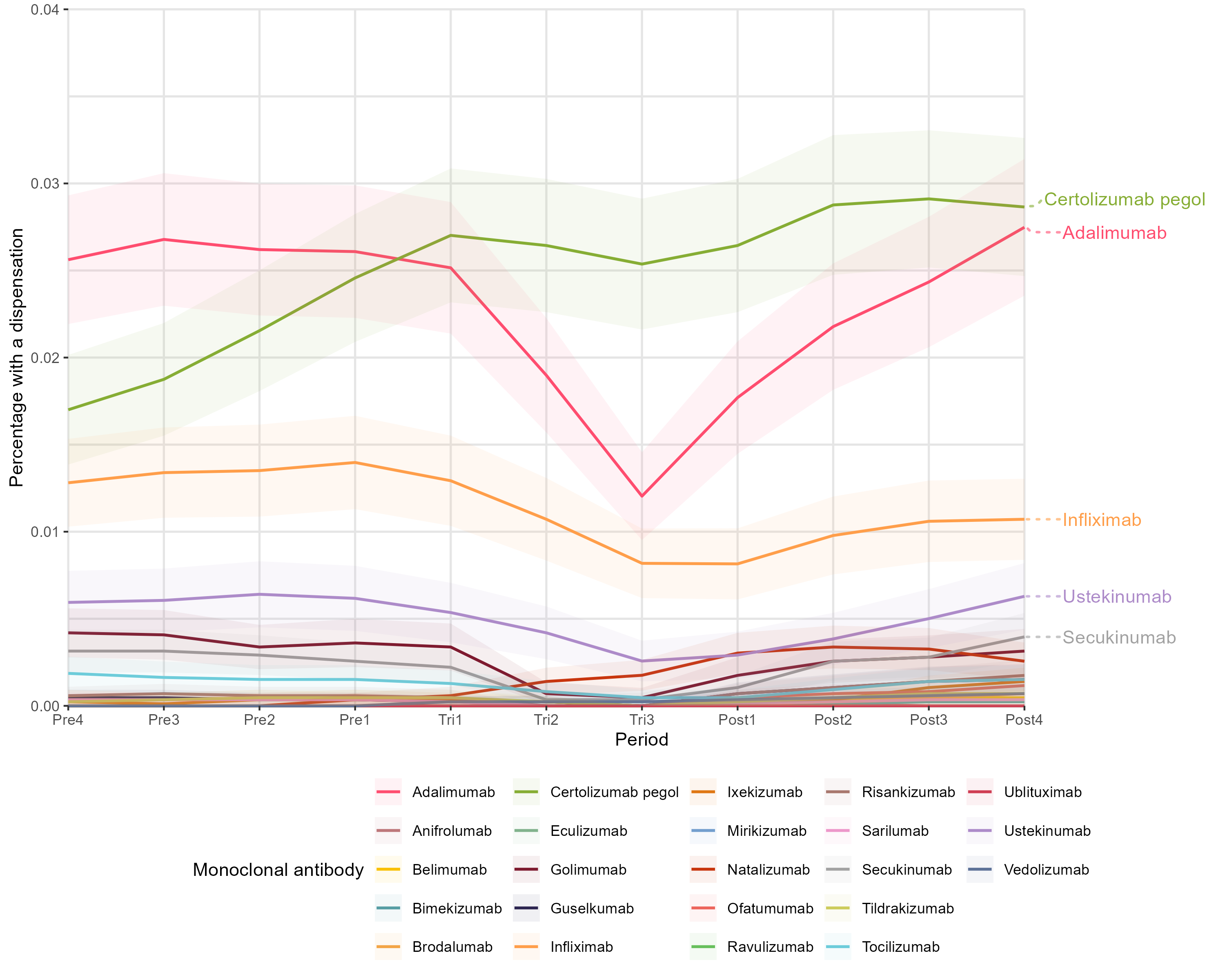

#### Figure S10. Prevalence of monoclonal antibody prescribing during and around pregnancy by period, stratified by drug.

Bands illustrate 95% confidence intervals around annual prevalence estimates, accounting for multiple pregnancies to the same individual. Denominator is all pregnancies in the study period (2012-2024).

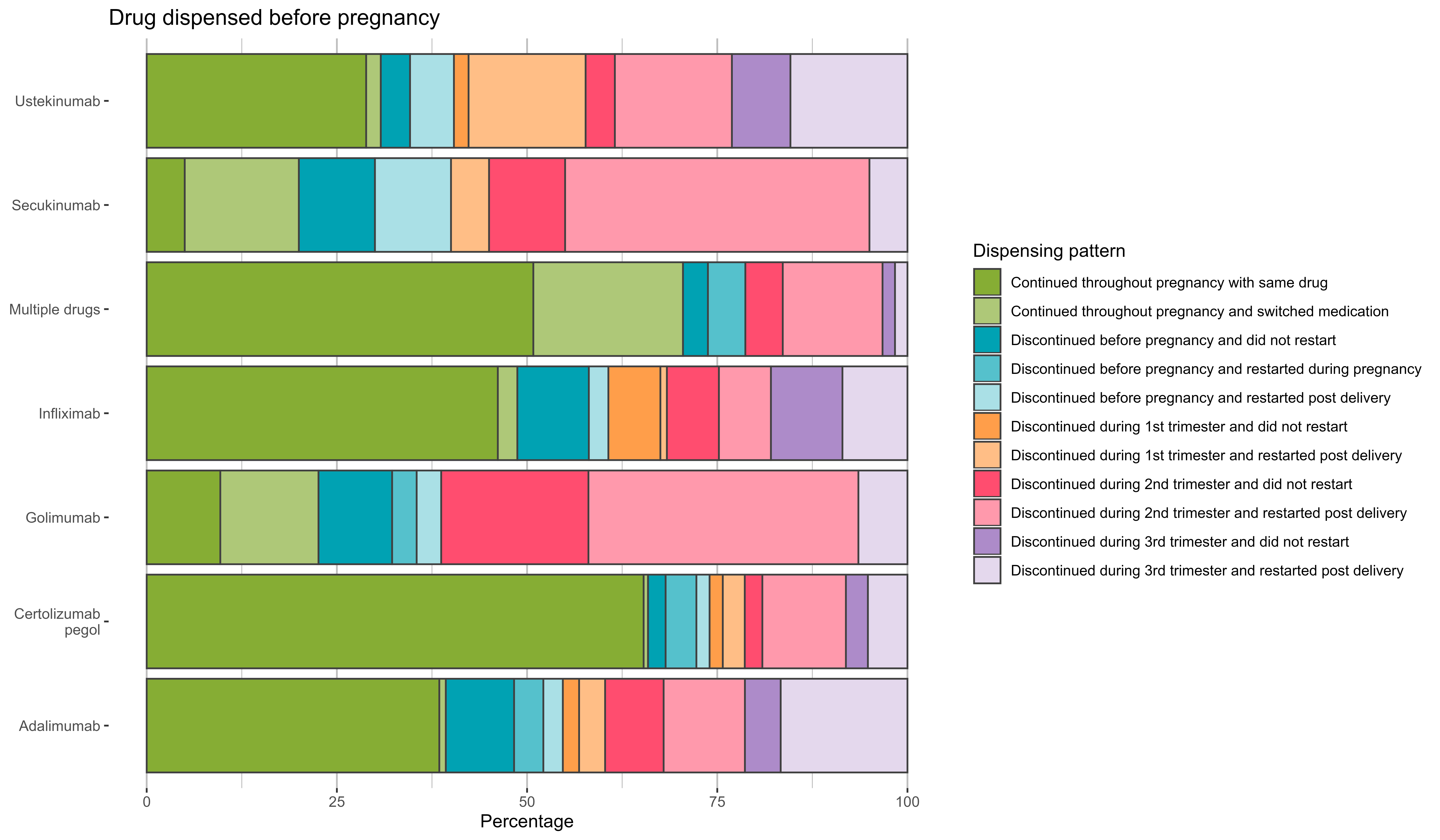

#### Figure S11. Prevalence of different dispensing patterns before and during pregnancy stratified by drug used in the year before pregnancy, among pre-existing users (n=719).

Pre-existing users defined as exposed to a mAb in at least one period between 1 year before and 90 days before pregnancy. Drugs used by fewer than 20 pregnancies were discarded in this analysis, resulting in a reduced sample size for this analysis (n=684/719 pre-existing users in sample).

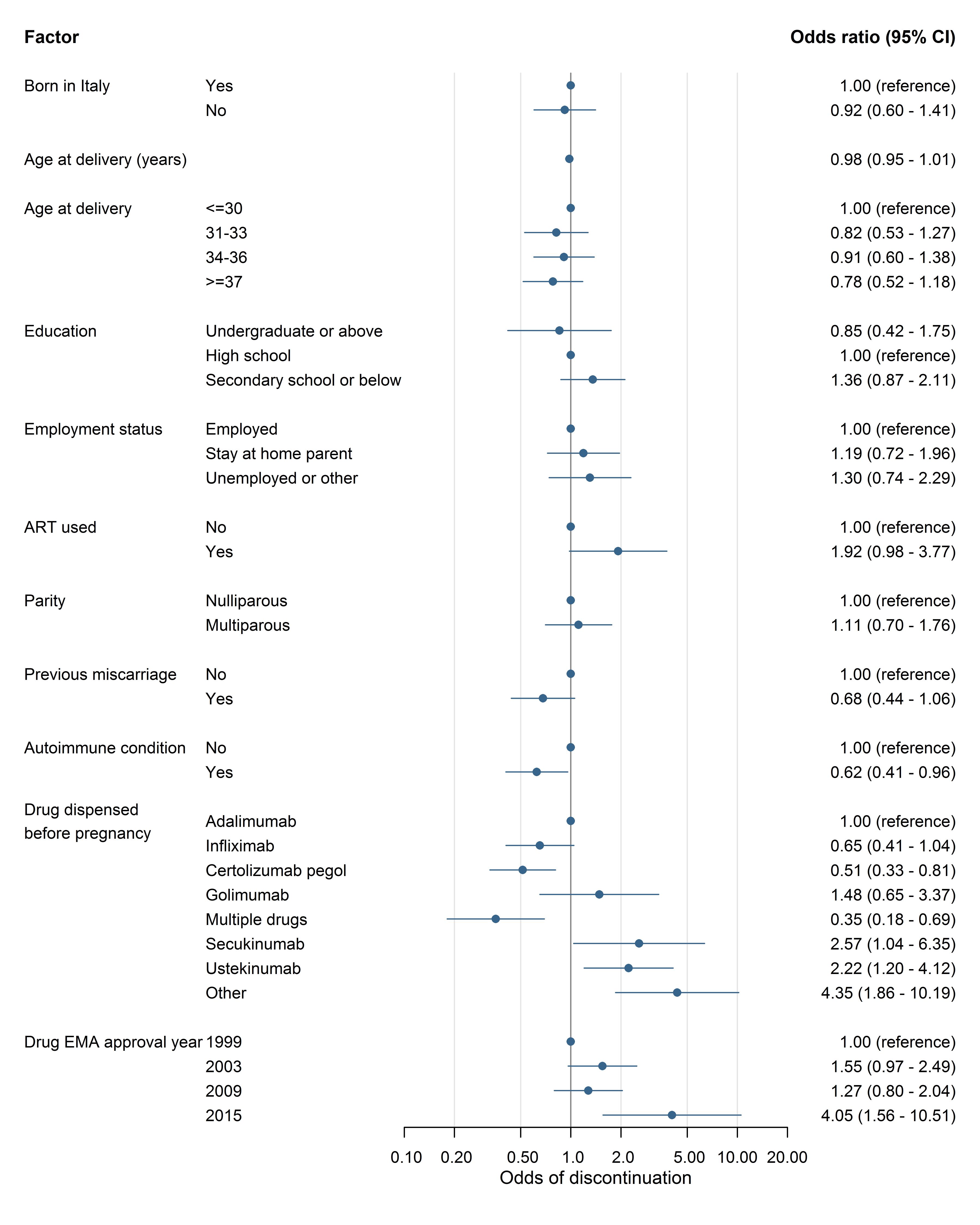

#### Figure S12. Associations between maternal factors and odds of discontinuation of mAb treatment during pregnancy, among pre-existing users (n=722).

All estimates were adjusted for the effect of calendar year only, and included complete records for each factor. Pre-existing user defined as exposed to a mAb between 1 year before and 90 days before pregnancy. ART = assistive reproductive technology. EMA = European Medicines Agency. Autoimmune condition diagnoses were derived from secondary care and/or dispensation payment exemption records only.

### Tables

#### Table S1. Key variables and the data sources.

These variables were used to stratify analyses and/or explored as factors which may be associated with discontinuation. ART = assisted reproductive technology. See Table S4 for codelists used to define each autoimmune condition.

| **Variable** | **Data source** | **Coding** |
| --- | --- | --- |
| Maternal age at delivery | Calculated from date of delivery (delivery records) and maternal birth year (demographic data) | Categoric:  <=30, 31-33, 34-36, >=37.  Continuous:  years |
| Country of birth | Demographic data | Categoric:  Born in Italy, Not born in Italy. |
| Maternal education | Delivery records | Categoric:  Secondary school or below, High school, Undergraduate, Postgraduate. |
| Calendar year of delivery | Delivery records | Categoric:  2012-14, 2015-17, 2018-20, 2021-24.  Continuous:  years |
| Use of ART | Delivery records | Binary:  Yes, no. |
| Drug dispensed in year before pregnancy | Pregnancy period calculated from date of delivery and gestational age at birth, both from delivery records.  Drug dispensations from hospital dispensation data, covering the year before pregnancy started. | Categoric:  Adalimumab, Infliximab, Certolizumab pegol, Golimumab, Multiple drugs, Secukinumab, Ustekinumab, Other (for drugs used before <20 pregnancies). |
| Drug EMA approval year | Drug obtained from ‘Drug dispensed in year before pregnancy’ variable. EMA for approval year (retrieved 20/02/2026 (4)). | Categoric:  1999 (Infliximab), 2003 (Adalimumab), 2009 (Certolizumab pegol, Ustekinumab, Golimumab), 2015 (Secukinumab). |
| Employment status | Delivery records | Categoric:  Employed, Stay at home parent, Unemployed or other (Unemployed, Looking for first job, Student or Other (retired, unable)). |
| Autoimmune condition | Outpatient hospital admissions, emergency room admissions, reasons for payment exemptions. | Binary:  Any recorded diagnosis prior to pregnancy or any payment exemption, or none.  Categoric:  Inflammatory bowel disease, Rheumatoid arthritis, Ankylosing spondylitis, Psoriasis, Multiple sclerosis, Systemic lupus erythematosus, Myasthenia gravis. |
| Parity | Delivery records | Binary:  Nulliparous, multiparous. |
| Previous miscarriage | Delivery records | Binary:  Yes, no. |

#### Table S2. Monoclonal antibodies used to manage autoimmune conditions which may be diagnosed in women of reproductive age (n=23 drugs).

ATC = Anatomical Therapeutic Chemical classification system.

| **Drug name** | **ATC code** | **Target** | **Autoimmune indications** |
| --- | --- | --- | --- |
| Ravulizumab | L04AJ02 | C5 | Myasthenia gravis |
| Eculizumab | L04AJ01 | C5 | Myasthenia gravis |
| Ublituximab | L04AG14 | MS4A1 | Multiple sclerosis |
| Ofatumumab | L04AG12 | MS4A1 | Multiple sclerosis |
| Anifrolumab | L04AG11 | IFNAR1 | Systemic lupus erythematosus |
| Vedolizumab | L04AG05 | ITGA4; ITGB7 | Inflammatory bowel disease |
| Belimumab | L04AG04 | TNFSF13B | Systemic lupus erythematosus |
| Natalizumab | L04AG03 | ITGA4 | Multiple sclerosis; Inflammatory bowel disease (Crohn's disease) |
| Mirikizumab | L04AC24 | IL23A | Inflammatory bowel disease |
| Bimekizumab | L04AC21 | IL17A; IL17F | Psoriasis; Ankylosing spondylitis |
| Risankizumab | L04AC18 | IL12B; IL23A | Psoriasis; Inflammatory bowel disease |
| Tildrakizumab | L04AC17 | IL23A | Psoriasis |
| Guselkumab | L04AC16 | IL23A | Psoriasis |
| Sarilumab | L04AC14 | IL6R | Rheumatoid arthritis |
| Ixekizumab | L04AC13 | IL17A | Psoriasis; Ankylosing spondylitis |
| Brodalumab | L04AC12 | IL17RA | Psoriasis |
| Secukinumab | L04AC10 | IL17A | Psoriasis |
| Tocilizumab | L04AC07 | IL6R | Rheumatoid arthritis |
| Ustekinumab | L04AC05 | IL12B | Psoriasis; Inflammatory bowel disease |
| Golimumab | L04AB06 | TNF | Rheumatoid arthritis; Psoriasis (psoriatic arthritis) ; Ankylosing spondylitis; Inflammatory bowel disease |
| Certolizumab pegol | L04AB05 | TNF | Rheumatoid arthritis; Psoriasis (psoriatic arthritis) ; Ankylosing spondylitis; Inflammatory bowel disease; Psoriasis |
| Adalimumab | L04AB04 | TNF | Rheumatoid arthritis; Psoriasis (psoriatic arthritis) ; Ankylosing spondylitis; Inflammatory bowel disease; Psoriasis |
| Infliximab | L04AB02 | TNF | Rheumatoid arthritis; Ankylosing spondylitis; Inflammatory bowel disease |

#### Table S3. Definitions for time periods of study across the year before pregnancy, during pregnancy, and the year following pregnancy.

| **Period** | **Start (inclusive)** | **End (inclusive)** |
| --- | --- | --- |
| 9-12 months pre-pregnancy | conception - 1 year | conception - 271 days |
| 6-9 months pre-pregnancy | conception - 270 days | conception - 181 days |
| 3-6 months pre-pregnancy | conception - 180 days | conception - 91 days |
| 0-3 months pre-pregnancy | conception - 90 days | conception - 1 days |
| 1st trimester | conception | conception + 90 days |
| 2nd trimester | conception + 91 days | conception + 180 days |
| 3rd trimester | conception + 181 days | delivery |
| 0-3 months postpartum | delivery + 1 day | delivery + 90 days |
| 3-6 months postpartum | delivery + 91 days | delivery + 180 days |
| 6-9 months postpartum | delivery + 181 days | delivery + 270 days |
| 9-12 months postpartum | delivery + 271 days | delivery + 1 year |

#### Table S4. ICD-9 codelists and prescription payment exemption codes from Regione Lombardia used to identify autoimmune conditions.

Diagnoses were obtained from hospital admissions and/or emergency room admissions.

| **Condition** | **ICD-9 codes** | **Payment exemption codes** |
| --- | --- | --- |
| Inflammatory bowel disease | 555*, 556* | 009.555, 009.556 |
| Rheumatoid arthritis | 714 | 006.714.0, 006.714.2 |
| Ankylosing spondylitis | 7200 | 054.720.0 |
| Psoriasis | 6960, 6961, 6968 | 045.696.0, 045.696.1 |
| Multiple sclerosis | 340 | 046.340 |
| Systemic lupus erythematosus | 7100 | 028.710.0 |
| Myasthenia gravis | 358 | 034.558.0, RFG101 |

#### Table S5. Counts of pregnancies following dispensing patterns, among all pregnant women with a recorded mAb prescription in at least one period before pregnancy (pre-existing users, n=722).

| **Dispensing pattern** | **Count (%)** |
| --- | --- |
| Continued throughout pregnancy and switched medication | 28 (3.9%) |
| Continued throughout pregnancy with same drug | 309 (42.8%) |
| Discontinued before pregnancy and did not restart | 49 (6.8%) |
| Discontinued before pregnancy and restarted during pregnancy | 20 (2.8%) |
| Discontinued before pregnancy and restarted post delivery | 20 (2.8%) |
| Discontinued during 1^st^ trimester and did not restart | 19 (2.6%) |
| Discontinued during 1^st^ trimester and restarted post delivery | 26 (3.6%) |
| Discontinued during 2^nd^ trimester and did not restart | 48 (6.6%) |
| Discontinued during 2^nd^ trimester and restarted post delivery | 97 (13.4%) |
| Discontinued during 3^rd^ trimester and did not restart | 34 (4.7%) |
| Discontinued during 3^rd^ trimester and restarted post delivery | 72 (10.0%) |
